# Anticoagulation and associated complications in Veno-Arterial Extracorporeal Membrane Oxygenation in Adult Patients: A Protocol for a Systematic Review and Meta-Analysis

**DOI:** 10.1101/2023.04.06.23288275

**Authors:** Ruan Vlok, Hergen Buscher, Anthony Delaney, Tessa Garside, John Myburgh, Priya Nair

## Abstract

**Background:** Veno-arterial Extracorporeal Membrane Oxygenation (VA-ECMO) is a rapidly expanding therapy with a relatively limited evidence base. Due to both quantitative and qualitative acquired coagulopathies in VA-ECMO, bleeding remains a major complication and with potentially catastrophic outcomes. Simultaneously, coagulation activation occurs via blood contact with the extracorporeal circuit, which risks not only vital organ emboli, but also the circuit viability. This carries the risk of equally catastrophic complications. As such, VA-ECMO patients are routinely anticoagulated. A large variation in practice exists surrounding anticoagulation in VA-ECMO. Despite an increasing uptake in the therapy, the evidence base is limited and current guidelines surrounding anticoagulation practice are based on expert opinion. We will perform a study level systematic review and meta-analysis of VA-ECMO patients comparing anticoagulation strategies, including the agent of anticoagulation, the method of monitoring and the targeted degree of anticoagulation to assess the association between described anticoagulation strategies and bleeding and thrombotic complications.

**Methods:** We will perform a study level meta-analysis of randomised controlled trials (RCTs) and cohort studies that report on bleeding and thrombotic complications in VA-ECMO. Our primary objective is to describe the incidence of bleeding and thrombotic complications associated with individual anticoagulation strategies. In order to be included, a study must report the anticoagulant drug used, the method of monitoring used, or the anticoagulation target used, and at least one outcome of interest. Four databases will be systematically reviewed by two authors. Two authors will extract and assess for risk of bias using the Cochrane Risk of Bias tool for RCTs and the ROBINS-I tool for observational studies. Data will be analysed via incidence rates of bleeding and thrombotic complications, with a subgroup analysis of each anticoagulation strategy where able. The study will be reported in line with PRISMA guidelines.

## BACKGROUND

Extracorporeal Membrane Oxygenation (ECMO) is a therapy that is rapidly expanding in intensive care practice as a rescue strategy for both cardiovascular and respiratory failure. Although ECMO is widely employed, it has limited high quality evidence to define best practice. As ECMO involves contact of blood with an extracorporeal pump, membrane, and circuit, anticoagulation is routinely used to maintain circuit patency and protect against thrombotic complications. In VA-ECMO low flow in the cardio-pulmonary vasculature may increase the thromboembolic risk. Both bleeding and thrombosis are associated with increased mortality in ECMO [1-3]. The optimal anticoagulation strategy in ECMO remains under debate and may require tailoring to individual risk factors. Modifiable factors include the dose and drug used for anticoagulation as well as the modality of monitoring and titrating it. Anticoagulation strategies include full dose therapeutic anticoagulation, low dose anticoagulation and no anticoagulation [4,5]. Anticoagulants described in the literature include unfractionated heparin, bivalirudin and argatroban [6,7]. Options for monitoring anticoagulation include activated partial prothrombin time (aPTT), anti-Xa assays, and point of care tests such as ACT or viscoelastic testing [8, 9].

Bleeding is a common complication, particularly of VA-ECMO. A recent meta-analysis described mean of 2.6 units of packed red cells transfused per day in ECMO patients, with veno-arterial (VA) modes requiring more transfusions on average compared to veno-venous (VV) [10]. Bleeding occurs as a result of not only anticoagulation but also acquired coagulopathy. Coagulopathy in ECMO may include thrombocytopenia, qualitative platelet dysfunction, coagulation factor consumption, hyperfibrinolysis and acquired von Willebrand factor dysfunction [11,12]. The quantitative and qualitative nature of the coagulopathy has driven discussion about the ideal monitoring tests [13]. The acquired coagulopathy differs between VV and VA-ECMO [11]. Bleeding may manifest as cannula site bleeding, surgical site bleeding, gastrointestinal bleeding and most devastatingly intracranial bleeding. The total packed red cell transfusion requirement on has been associated with increased mortality [2]. Conversely, thrombotic complications may be equally catastrophic. These include compromise of the circuit and oxygenator which may require routine or emergent circuit change, as well as emboli to vital organs and deep vein thrombosis.

Potential interventions for minimising complications associated with anticoagulation include anticoagulant selection, anticoagulant dosing, monitoring, cannulation strategies and correction of coagulopathy. The Extracorporeal Life Support Organization (ELSO) recommends using unfractionated heparin, targeting an activated clotting time ACT of 180-220 seconds based on limited data and expert opinion [14]. In order to define the optimal strategy to minimise complications associated with anticoagulation, risk factors need to be profiled. Antithrombin III levels have been shown to be lower in VA compared to VV ECMO and VA-ECMO patients have been described as spending less time in therapeutic ranges when using unfractionated heparin, implying the strategies may differ between VA and VV-ECMO patients [11]. Differences in coagulopathy appear to exist within VA patients as well, with more hypofibrinogenaemia proposed to exist in extracorporeal cardiopulmonary resuscitation (E-CPR) patients [11].

We will perform a study level systematic review and meta-analysis of VA-ECMO patients comparing anticoagulation strategies, that is the agent of anticoagulation, the method of monitoring and the targeted degree of anticoagulation to assess the association between described anticoagulation strategies and bleeding and thrombotic complications. This systematic review and meta-analysis will have an exploratory role, given the anticipated limited available literature and wide variation in clinical care of ECMO patients outside of anticoagulation strategies, and will not aim to describe a single best anticoagulation strategy.

### OBJECTIVES

We will perform a systematic review and meta-analysis describing the association between anticoagulation strategy of choice and the incidence of complications associated with anticoagulation in VA-ECMO. We will also aim to describe the features that predispose to or protect from these complications. We will report according to the PRISMA guidelines [15] and will publish the protocol on a preprint server [16]. Our primary objective is to describe the incidence of bleeding and thrombotic complications associated with individual anticoagulation strategies described in the published literature. Due to the expected heterogeneity, we aim to assess the association between anticoagulant drug choices, anticoagulant dosing and anticoagulant monitoring, as well as clinical features such as ECPR, post cardiotomy ECMO and cannula configuration and haemostatic complications.

## METHODS

### 1. Criteria for considering studies for this review

#### 1.A Types of studies

We will include all randomized controlled trials (RCTs), quasi-RCTs and controlled and uncontrolled cohort studies that describe anticoagulation and bleeding or thrombotic complications in adult patients on VA-ECMO. We define quasi-randomization as non-true randomization to each arm by means such as alternation, case record number or date of birth. Case series and cohort studies with a sample less than ten patients will be excluded. We will exclude physiological modelling studies or case reports. Studies that include both VA and VV-ECMO will be included if the VA patients’ results are reported separately. There will be no language restrictions to included studies. Abstracts and studies in press will be included. Due to the evolution in ECMO technology and accumulating institutional experience, only studies published after 2010 will be included.

#### 1.B Types of participants

We will include studies involving adults aged 16 years and older admitted to intensive care for any indication who were commenced on VA-ECMO.

#### 1.C Types of interventions

We will include studies that assess any intervention in patients on VA-ECMO. All cannulation configurations will be included, including hybrid configurations, central and peripheral cannulation will be included. Veno-pulmonary arterial ECMO (V-PA) will not be considered VA ECMO and will not be included. All indications for ECMO will be included, including post-cardiotomy and extracorporeal cardiopulmonary resuscitation (E-CPR). Where studies report a mixed cohort of VV and VA-ECMO, only studies that describe outcomes of interest separately for the VA-ECMO patients will be included. In order to be included, a study must report the following:

1. the anticoagulant drug used OR
2. the test for monitoring used AND
3. at least 1 included outcome of interest.

### 2. Data collection and analysis

#### 2.A Selection of studies

Two review authors (RV and one additional author) will independently screen all titles and abstracts of each reference identified by our search and will then independently assess the full text of any potentially relevant studies for eligibility or exclusion. We will use Covidence software to collate search results, remove duplicates and record screening decisions and exclusions at each stage [17]. This will be facilitated by a standardized digital screening, inclusion and exclusion tools, in line with methods outlined in the *Cochrane Handbook for Systematic Reviews of Interventions* [18]. We will resolve any disagreements by discussion and consensus prior to proceeding at each stage.

We will record the process in sufficient detail to produce a PRISMA flow diagram [15]. For all full-text articles screened and excluded, we will record the reason for exclusion in sufficient detail to present a table of characteristics for excluded studies.

#### 2.B Data extraction and management

Through Covidence, we will use a standardized digital data-extraction sheet, in line with requirements outlined in the *Cochrane Handbook for Systematic Reviews of Interventions Version 6*.*3* [18]. Two review authors (RV and one additional author) will independently extract information regarding trial design, data regarding the anticoagulation strategy, outcomes of interest (as reported below) and information relevant to Risk of Bias grading. Where required, we (RV and one additional author) will contact individual trial authors or organizations to obtain missing data or clarification regarding unclear data. We will resolve any disagreements by discussion and consensus.

#### 2.C Assessment of risk of bias in included studies

Two review authors (RV and one additional author) will independently assess methodological quality. For all included trials we will utilize Cochrane’s Risk of Bias 2 (ROB 2) tool for RCTs [18]. There are five specific measurements covered by this tool, and the *Cochrane Handbook for Systematic Reviews of Interventions* describes how each domain should be assessed to reach a judgement of low risk, some concerns or high risk of bias. If a study is judged to be low risk in all domains, then it will be deemed to be at low risk of bias overall.

The criteria include:

1. Bias arising from randomisation process;
2. Biase due to deviations from intended intervention;
3. Bias due to missing outcome data;
4. Bias in measurement of outcome data;
5. Bias in selection of the reported data;

For non-randomized controlled trials, the ROBINS-I tool will be used to assess risk of bias [19]. The criteria for the ROBINS-I tool include pre-intervention, at intervention and post-intervention domains with assessment of selection bias, information bias, confounding and reporting bias. Bias surrounding deviation in intended intervention is anticipated, given observational studies are unlikely to report on intentional deviation of anticoagulation management from institutional protocols in patients with increased bleeding or thrombosis.

We will present a ‘Risk of Bias’ summary, as well as ‘Risk of Bias’ judgements for individual studies in the ‘Characteristics of included studies’ tables. Given anticipated heterogeneity in patients and clinical practice surrounding management of VA-ECMO, we will be hesitant drawing firm conclusions from the pooled analyses.

### 2. Important outcomes

Unless otherwise specified, outcomes will be limited to the duration of the time spent on ECMO.

1. Primary Outcomes:
  A. Bleeding events-A composite outcome of Major Bleeding Events (as defined by the study), and intracranial bleeding. For intracranial bleeding to be included in the composite, the events should not have been included in the definition of Major Bleeding Events.
  B. Thrombotic events - A composite outcome of major thromboembolic events as defined by the study, thromboembolic stroke or distal organ emboli and circuit or circuit component exchange
2. Secondary Bleeding Outcomes
  A. Cannula site bleeding
  B. Gastrointestinal, intraabdominal and retroperitoneal bleeding
  C. Intracranial bleeding
  D. Respiratory tract bleeding (pulmonary, tracheobronchial and oropharyngeal)
  E. Thoracic bleeding
  F. Bleeding prompting need for procedural intervention (surgery, endoscopy, interventional radiology or new drain insertion)
  G. Requirement for Massive Transfusion
  H. Bleeding definition used
3. Secondary Thrombotic outcomes
  a. Circuit or component change
  b. Deep venous thrombosis (if specifically described, DVTs associated with cannulation sites post decannulation will be included)
  c. Thromboembolic ischaemic stroke
4. Survival to decannulation
5. Duration of ECMO run
6. Mortality at Longest Follow-Up

### 3. Search methods for identification of studies

#### 4.A Electronic searches

We will search electronic databases for articles using key words, synonyms and subject headings that relate to ECMO, anticoagulation and relevant complications. We will use controlled vocabulary specific to each database (see ‘Search Strategy’). Two review authors (RV and one additional author) will perform the search independently and no language restrictions will be applied. We will contact individual trial authors for additional information where necessary.

We will search the following databases for published trials:

1. Cochrane Central Register of Controlled Trials (CENTRAL) (1996 to present)
2. MEDLINE Ovid (1946 to present)
3. Elsevier Embase (1947 to present)
4. CINAHL EBSCO (1937 to present)

#### 4.B Searching other resources

We will perform a citation search of all included studies as well as any relevant studies and reviews on anticoagulation for ECMO. If this identifies any additional eligible studies, we will re-examine and update the search strategy. We will also perform a search of Google Scholar for additional trials. We will consider for inclusion studies that are available as abstracts only.

### 5. Data synthesis

Analyses will be performed using STATA (MP version 16.1, College Station, TX, USA). Separate analyses will be performed for controlled and non-controlled studies. It is anticipated that most studies will be single arm cohort studies and as such, analysis will be limited in these cases to incidence rates. For controlled studies, risk ratios and mean differences will be reported. Incidence rates of mortality, bleeding events, and thromboembolic events will be calculated by pooling study-specific data. We will use a random effects meta-analysis model using the DerSimonian and Laird method for proportions using the metaprop_one command in STATA, with 95% confidence intervals. Estimates will be represented in forest plots. Where possible, sensitivity a sensitivity analysis will be performed to assess high quality studies. Publication bias will be assessed by funnel plot and Egger test. Where able, for dichotomous outcomes, we will calculate risk ratios (ORs) with 95% confidence intervals (CIs). For continuous outcomes, we will calculate mean differences (MDs) with 95% CIs. When appropriate, we plan to calculate a number needed to treat for an additional beneficial outcome (NNTB) or a number needed to treat for an additional harmful outcome (NNTH) to aid clinical decision making.

#### 5.A Subgroup analysis and investigation of heterogeneity

Where sufficient data is available, subgroup analyses will be performed for each outcome based on:

1. Anticoagulant choice
2. Anticoagulation target
3. Method of monitoring anticoagulation
4. Cannula configuration (central vs peripheral, percutaneous vs open peripheral)
5. ECPR
6. Post cardiotomy
7. Oxygenator surface area (=<2.0 m^2^)

#### 5.B Sensitivity analysis

We will perform a sensitivity analysis of studies to explore the influence of ‘Risk of bias’ assessments. This will allow assessment of the robustness of our conclusions. These sensitivity analyses will be limited to the primary outcomes of composite major bleeding events and composite major thrombotic events.

#### 5.C Dealing with missing data

We will contact trial authors to request missing data. We will outline any missing data in the results and explain in the discussion the extent to which the overall results may be affected. In the event of unreported standard deviations, we will make an estimate based on other reports within the meta-analysis.

#### 5.D Assessment of heterogeneity

If there is obvious extensive clinical heterogeneity, we will not perform a meta-analysis. Otherwise, we will investigate clinical heterogeneity via meta-regression, identifying trends within subgroups of the clinical population as detailed above.

We will utilize the I^2^ test to assess statistical heterogeneity for each outcome [17]. We will judge I^2^ values as follows;

- 0% to 40%: might not be important;
- 41% to 60%: may represent moderate heterogeneity.
- 61% to 90%: may represent substantial heterogeneity.

## Intended Search Strategy in Ovid Format

Database: Ovid MEDLINE(R) ALL <1946 to Present>

Search Strategy:

1. ECMO.tw. or Extracorporeal Membrane Oxygenation/
2. (ECLS or Extracorporeal Life Support).tw.
3. extracorporeal.tw. or Extracorporeal Circulation/
4. ‘Extracorporeal cardiopulmonary resuscitation’.tw.
5. ECPR.tw.
6. 1 or 2 or 3 or 4 or 5
7. Anticoagula*.tw. or Anticoagulants/
8. (heparin or UFH or unfractionated).tw. or Heparin/
9. bivalirudin.tw.
10. Nafamosta.tw.
11. prostacyclin.tw.
12. epoprostenol .tw.
13. argatroban.tw.
14. APTT.tw. or Partial Thromboplastin Time/ or Blood Coagulation/ or Blood Coagulation Tests/ or Prothrombin Time/
15. (TEG or thromboelastography).tw.
16. (ROTEM or thromboelastometry).tw.
17. viscoelastic.tw. (16726)
18. (activated clotting time or ACT).tw.
19. Anti$XA.tw.
20. Platelet Function Tests/
21. 7 or 8 or 9 or 10 or 11 or 12 or 13 or 14 or 15 or 16 or 17 or 18 or 19 or 20
22. Bleed*.tw.
23. Gastrointestinal Hemorrhage/
24. Hemostasis/ or h*emosta*.tw.
25. Postoperative Complications/ or Postoperative Hemorrhage/ or Reoperation/
26. (‘return to theatre’ or ‘rethoracotomy’).tw.
27. Thrombosis/ or thromb*.tw.
28. Intracranial Thrombosis/ or Embolic Stroke/ or Hemorrhagic Stroke/ or Ischemic Stroke/ or Stroke/ or Thrombotic Stroke/
29. Venous Thrombosis/ or Pulmonary Embolism/ or Venous Thromboembolism/
30. (DVT or deep vein thrombosis).tw.
31. Ischemia/ or Peripheral Arterial Disease/ or Arterial Occlusive Diseases/
32. Blood Component Transfusion/ or Blood Transfusion/ or Erythrocyte Transfusion/ or Platelet Transfusion/ (69654)
33. transfus*.tw.
34. (‘massive transfusion’ or MTP).tw.
35. ‘bleeding event’.tw.
36. ‘thrombotic event’.tw.
37. BARC.tw.
38. Thromb$s.tw.
39. 22 or 23 or 24 or 25 or 26 or 27 or 28 or 29 or 30 or 31 or 32 or 33 or 34 or 35 or 36 or 37 or 38
40. 6 and 21 and 39

## Data Availability

This manuscript is a protocol for a systematic review

